# Data Resource Profile: EST-Health-30

**DOI:** 10.64898/2026.04.21.26351087

**Authors:** Sulev Reisberg, Marek Oja, Kerli Mooses, Sirli Tamm, Ami Sild, Harry-Anton Talvik, Sven Laur, Raivo Kolde, Jaak Vilo

## Abstract

**Background:** The increasing availability of routinely collected health data offers new opportunities for population-level research, yet access to comprehensive, linked, and standardised datasets remains limited. We describe EST-Health-30, a large-scale, population-representative health data resource from Estonia.

**Methods:** EST-Health-30 comprises a random 30% sample of the Estonian population (~500,000 individuals), with longitudinal data from 2012 to 2024 and annual updates planned through 2026.Individual-level records are linked across five nationwide databases, including electronic health records, health insurance claims, prescription data, cancer registry, and cause of death records. A privacy-preserving hashing approach ensures consistent cohort inclusion over time while maintaining pseudonymisation. All data are harmonised to the Observational Medical Outcomes Partnership (OMOP) Common Data Model (version 5.4) using international standard vocabularies. Data quality was assessed using established OMOP-based validation frameworks.

**Results:** The dataset contains rich multimodal information on diagnoses, procedures, laboratory measurements, prescriptions, free-text clinical notes, healthcare utilisation, and costs, with high population coverage and longitudinal depth. Data quality assessment showed high completeness and consistency, with 99.2% of applicable checks passing. The age–sex distribution closely reflects the national population, supporting representativeness, though coverage is marginally below the target 30% (29.2%), primarily attributable to recent immigrants without health system contact. The dataset enables construction of detailed clinical cohorts, analysis of disease trajectories, and evaluation of healthcare utilisation and outcomes across the life course.

**Conclusions:** EST-Health-30 is a comprehensive, standardised, and population-representative real-world data resource that supports epidemiological, clinical, and methodological research. Its alignment with the OMOP CDM facilitates reproducible analytics and participation in international federated research networks, while secure access infrastructure ensures compliance with data protection regulations.

**Key features:**

- EST-Health-30 is a population-representative dataset of complete health records for a random 30% sample of the Estonian population (~500,000 individuals) spanning 2012-present, enabling population-level epidemiological analyses with annual updates.
- The dataset is constructed using a random sampling approach based on hashed password-protected personal identifiers, ensuring consistent inclusion over time with unbiased population coverage.
- Individual-level data are linked across multiple nationwide databases, including electronic health records, claims, prescriptions, cancer and cause of death registry data, enabling multimodal analyses of health trajectories.
- All data are standardised to the OMOP Common Data Model (CDM) version 5.4 using international vocabularies (e.g., SNOMED CT, RxNorm, LOINC), supporting reproducibility and participation in federated research networks.
- The dataset is accessible through a secure processing environment compliant with the European Health Data Space (EHDS) framework.

## Data resource basics

EST-Health-30 is a population-based health dataset that includes complete health records for a randomly sampled 30% of the Estonian population, linked across five national databases: electronic health records from primary and secondary care, claims, laboratory test results, pharmacy data, and cancer and cause of death registries, from 2012 onwards. The dataset currently contains data through 2024 and includes 509,856 individuals.

It is updated annually through 2026, with each release typically including data up to the end of the previous calendar year. Due to processing and validation procedures, the expected lag between data generation and availability is approximately 6–24 months, depending on the source (e.g., manually curated cancer registry data may have longer delays). The dataset is fully mapped to the Observational Medical Outcomes Partnership (OMOP) Common Data Model (CDM), enabling a wide range of study designs.

## Data collected

### Source data

#### Source population

Estonia is a Northern European country on the Baltic Sea with a population of about 1.37 million, of whom about 70% are ethnic Estonians and 82% hold Estonian citizenship. Most of the population speaks Estonian as their first language. Although the country has experienced a negative natural population growth since the 1990s, its population size has remained relatively stable since 2000 due to positive net migration. However, there was a spike in 2022 due to Ukrainian refugees, when the net migration was nearly 40,000 people compared to 5,700, an average net migration during five consecutive years before, increasing the population size by 2.6%. In 2024, 9,690 births and 15,756 deaths were recorded. The life expectancy for those born in 2024 is 79.5 years (75.0 years for men and 83.4 years for women), which is slightly less than the average in the European Union (81.7 years) ^1,2^.

#### Estonian healthcare system

A comprehensive overview of the Estonian healthcare system is provided by Kasekamp et al. ^3^. Around 94% of the population is insured through the state-funded system managed by the Estonian Health Insurance Fund (EHIF). Roughly half of those insured are covered via employment-based or state contributions, while the remainder — such as children and individuals unable to work — receive coverage without contributing directly. EHIF finances a broad range of healthcare services, and even those without insurance have access to emergency care, cancer screening, treatment for HIV and tuberculosis, and smoking or addiction cessation programs. Beyond the primary care system, which is based on family doctors, patients typically pay in full or share costs for dental services (except for children under 19), long-term care, and many medications, particularly over-the-counter ones. In 2023, Estonia had the highest rate of unmet need for medical examination in the EU (12.9% vs EU average of 2.4%), with waiting lists as the primary driver ^3^. Because of long waiting times for some EHIF-funded specialist outpatient services, many patients choose to pay out-of-pocket for quicker access.

### Source datasets

Estonia is a leader in delivering essential public services through digital platforms ^4^. Regardless of whether individuals are insured, it is mandatory to collect and store health information in national databases for all individuals, resulting in longitudinal data. The EST-Health-30 dataset, compiled in 2025, integrates data from five national real-world health datasets (RWD) in Estonia:

1. E**stonian National Health Information System** (ENHIS), operational since 2008, is the backbone of the national health system. All officially recognized healthcare providers are legally required to submit patient case summaries to ENHIS, regardless of insurance coverage. The system collects primary and secondary care summaries (including both in- and outpatient records), referrals, referral responses (including laboratory results), and immunisation records as Health Level Seven (HL7) Clinical Document Architecture (CDA) documents. Case summaries include both structured and unstructured data. While EST-Health-30 contains the majority of the document types in ENHIS, it excludes certain types, such as medical images, dental care summaries, ambulance records, etc. Throughout this article, all documents retrieved from ENHIS are referred to as electronic health records (EHR).
2. **The Health Insurance Fund Database** — whose records we hereafter refer to as claims — contains information on diagnoses, procedures, claim costs, and insurance coverage periods, with data collected since the mid-1990s.
3. **Estonian Medical Prescription Center**, launched in 2010, replaced paper-based prescriptions. By 2014, 99% of prescriptions were electronic. Even paper prescriptions were entered into the digital system by pharmacies upon dispensing. The database excludes over-the-counter medications and drugs administered during hospital stays.
4. The **Cancer Registry**, operating since 1968, records all cancer cases in Estonia. Due to manual curation, data are available with an approximate two-year delay.
5. The **Causes of Death Register**, contains the information on all deaths in Estonia since 1986 ^5^.

An overview of the document types included in the EST-Health-30 datasets is given in **Table 1**.

**Table 1.** The summary statistics of the source documents in EST-Health-30 dataset.

| Document type | Content examples | Document count | Patients with at least one document (n) | Mean document count per patient (total cohort) |
| --- | --- | --- | --- | --- |
| Prescription | Prescription date, diagnosis (ICD-10), prescribed ingredients (ATC) and amount, dispensed package and amount, dispense date, costs for the patient and insurance fund | 40,028,909 | 467,858 | 78.5 |
| Claims: outpatient care | Visits, diagnoses (ICD-10), healthcare services provided, costs for the patient (for prescriptions only) and insurance fund | 29,143,369 | 488,114 | 57.2 |
| EHR: outpatient care | Visits, diagnoses (ICD-10), anamnesis, objective findings, allergies, lab/pathology/endoscopy/radiology results (except images), measurements, test results, procedures, surgeries, summary | 17,424,410 | 487,469 | 34.2 |
| EHR: referral response | Results of lab tests | 15,521,887 | 477,256 | 30.4 |
| Insurance period | Dates and the basis of the insurance period | 3,549,258 | 496,620 | 7.0 |
| EHR: immunisation | Immunization date and info | 2,132,845 | 349,446 | 4.2 |
| Claims: dental care | Same as ‘Claims: outpatient care’ | 1,747,102 | 165,580 | 3.4 |
| EHR: inpatient care | Same as ‘EHR: outpatient care’, length of hospital stay, drugs administered during the stay (inconsistently recorded) | 793,431 | 265,323 | 1.6 |
| Claims: inpatient care | Same as ‘Claims: outpatient care’ | 784,614 | 263,840 | 1.5 |
| Claims: nursing care | Same as ‘Claims: outpatient care’ | 330,627 | 45,301 | 0.65 |
| Claims: day care | Same as ‘Claims: outpatient care’ | 316,118 | 150,234 | 0.62 |
| EHR: day care | Same as ‘EHR: outpatient care’ | 243,349 | 125,350 | 0.48 |
| Death record | Death date, cause | 61,822 | 61,822 | 0.12 |
| Cancer registry record | Tumour diagnosis, topography, morphology, TNM classification, date, general info on first line treatment | 31,241 | 28,569 | 0.061 |
| EHR: home nursing | Visits, diagnoses (ICD-10), anthropometric measurements, information on smoking and alcohol/drug abuse, lifestyle, overview of health status | 8,141 | 5,326 | 0.016 |
| EHR: inpatient nursing | Same as ‘EHR: outpatient care’ | 2,957 | 2,314 | 0.006 |

Footnote: “Document count” refers to the number of source documents (e.g., Health Level Seven Clinical Document Architecture
documents or claims entries) rather than unique clinical encounters. “Mean document count per patient” reflects the average number
of documents per individual accumulated over the full observation period (2012–2024). Abbreviations: ICD-10, International
Classification of Diseases, 10th Revision; EHR, electronic health record.

## Data collection method

In the EST-Health-30 dataset, information from national health databases is linked at the individual level using a unique Personal ID—an immutable 11-digit identifier assigned to all Estonian citizens at birth and to permanent residents upon arrival. This identifier enables deterministic linkage of individual-level data across databases with high accuracy and consistency. The current dataset includes records from 2012 to 2024, with updates planned through 2026.

## Sampling

EST-Health-30 contains complete health records for a randomly selected 30% of the Estonian population, drawn from national databases. To generate this cohort, all data holders applied a shared SHA-256 hashing procedure to personal IDs padded with a secret key. Individuals whose hash values met predefined criteria — based on specific symbol positions — were included, ensuring exactly 30% random coverage. The resulting SHA-256 digests serve as pseudonyms in the dataset: they uniquely identify individuals within the dataset but cannot be used to recover the original personal identifiers.

For privacy protection, the only private information exchanged between database owners was a common secret key, used for hashing, as each party had knowledge of their own personal ID-s already. No raw data was shared, and the secret key remained unknown to the EST-Health-30 research team, preventing re-identification. The procedure allows data owners to process their data independently, while allowing the data recipient to link records by the same pseudonym (hash value), accounting for newly added individuals and maintaining consistent randomization over time.

**Figure 1** gives an overview of the population of the EST-Health-30 dataset, compared to the official Estonian population on 1st January 2024. In total, the resulting database contains health data of 509,856 individuals (244,659 males, 48.0%, and 265,095 females, 52.0%, and 102 with unspecified gender value, 0.02%), 401,272 of them observed on **1st of January 2024**.

**Figure 1.**
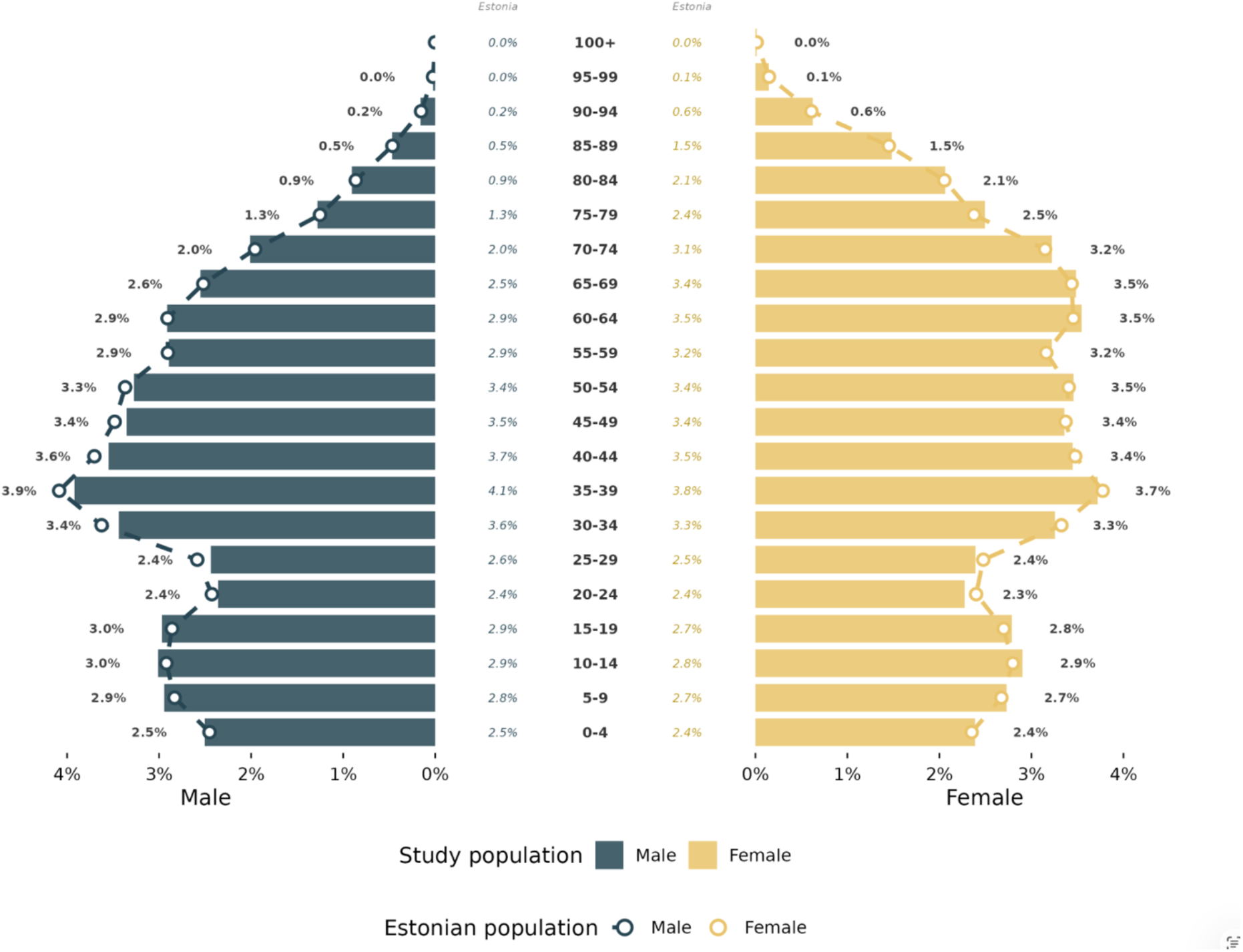
Population pyramid of the EST-Health-30 dataset (n=401,272), compared to official Estonian population (n=1,374,687) on 1st Jan 2024.

## Data harmonisation

All data sources are mapped to OMOP common data model (CDM) version 5.4 and its standard vocabularies by using the transformation pipeline described by Oja et al^6^. and Talvik et al^7^. This includes transforming original ICD-10 diagnosis codes and medication package codes to SNOMED CT and RxNorm. Source codes are retained alongside standardised concepts, allowing analyses to be performed using either representation depending on the use case. The following variables were also extracted from free-text fields written in Estonian and mapped to OMOP: body height, weight, body mass index, waist circumference, blood pressure, total-, LDL-, and HDL-cholesterol, estimated glomerular filtration rate (eGFR), left ventricular ejection fraction, prostate-specific antigen (PSA), Eastern Cooperative Oncology Group (ECOG) performance status, and tobacco smoking status. Free text parts from the EHR are stored in the notes table in OMOP CDM.

**Table 2** provides an overview of the main OMOP domains (tables) and their record counts in the resulting dataset. Since the dataset does not include migration records, observation periods were defined as follows. For each person, a candidate observation window was derived as the earliest of the first recorded clinical event and the start of their insurance period, through to the latest of the last recorded clinical event and the end of their insurance period. This window was then constrained by hard boundaries: the start date cannot precede 1 January 2012 or the person’s date of birth, and the end date cannot exceed 31 December 2024 or the person’s date of death, if recorded. For persons with neither clinical events nor an insurance period, a nominal one-day observation period of 1 January 2011 was assigned as a sentinel value; these are 6 persons who appear in the database solely because a drug was prescribed to them that was never dispensed. This sentinel value allows these records to be identified and excluded from analyses where a valid observation period is required. The lower mapping rate observed in the Procedure domain (84.3%) primarily reflects the complexity and heterogeneity of local procedure coding systems rather than deficiencies in data quality. Many local procedure codes are broad or ambiguous, covering multiple distinct procedures under a single code, and therefore do not have direct equivalents in standard vocabularies and require ongoing mapping efforts.

**Table 2.** Main domains (tables) and their features in the EST-Health-30 OMOP dataset.

| Domain | Type of available information | Record count | Person count | % Persons | Classifiers used (source and OMOP standard vocabulary) | Distinct source code count | Number of distinct source codes mapped to OMOP standard vocabularies | % Total codes mapped | % Total records mapped |
| --- | --- | --- | --- | --- | --- | --- | --- | --- | --- |
| Measurement | Date, value, unit, range | 248,358,784 | 493,663 | 96.8% | LOINC, Estonian local healthcare service codes, Cancer Modifier, OMOP Extension, SNOMED CT, ICD-10, SNOMED, NCSP | 5,874 | 4,776 | 81.3% | 97.4% |
| Cost | Total, paid by person and paid by insurance cost records for all domains | 166,179,250 | 494,709 | 97.3% | Euros | NA | NA | NA | NA |
| Note | Texts from EHR (in Estonian) | 115,251,935 | 496,071 | 97.3% | NA | NA | NA | NA | NA |
| Condition | Start and end date of the condition (mostly diagnosis), cause of death (if applicable) | 109,820,857 | 493,042 | 96.7% | SNOMED CT, ICD-10, ICD-O-3, OMOP Extension, NCSP, Estonian local healthcare service codes | 16,305 | 16,073 | 98.6% | 100.0% |
| Document (visit in OMOP) | Document type, date or case dates, care site | 108,477,759 | 509,849 | 100.0% | NA | 20 | 20 | 100.0% | 100.0% |
| Observation | Date, value & unit or qualifier | 86,673,044 | 493,502 | 96.8% | SNOMED CT, DRG, LOINC, OMOP Extension, Estonian local healthcare service codes, ICD-10, NCSP | 14,163 | 12,230 | 86.4% | 97.7% |
| Drug | Date of prescription and dispense, ingredient, package code, quantity, days supply | 41,992,793 | 476,413 | 93.4% | RxNorm, ATC, Estonian local drug package codes, CVX, Estonian local healthcare service codes, NCSP, SNOMED CT | 5,511 | 5,156 | 93.6% | 99.3% |
| Procedure | Date and procedure | 36,815,872 | 488,347 | 95.8% | SNOMED CT, Estonian local healthcare service codes, ICD-10, NCSP | 8,088 | 4,338 | 53.6% | 84.3% |
| Person | Birth year, gender, death date | 509,856 | 509,856 | 100.0% | NA | NA | NA | NA | NA |
| Observation period | Start and end date of the observation period of the person | 509,856 | 509,856 | 100.0% | NA | NA | NA | NA | NA |

Footnote: Abbreviations: ICD-10, International Classification of Diseases, 10th Revision; OMOP, Observational Medical Outcomes
Partnership; CDM, Common Data Model; SNOMED CT, Systematized Nomenclature of Medicine Clinical Terms; LOINC, Logical
Observation Identifiers Names and Codes; ATC, Anatomical Therapeutic Chemical classification system; NCSP, Nomesco Classification of
Surgical Procedures; ICD-O-3, The International Classification of Diseases for Oncology, 3rd Edition; DRG, Estonian diagnosis-related
groups; CVX, Vaccine Administered code set.

**Table 3** presents the ten most prevalent concepts within each domain.

**Table 3.**
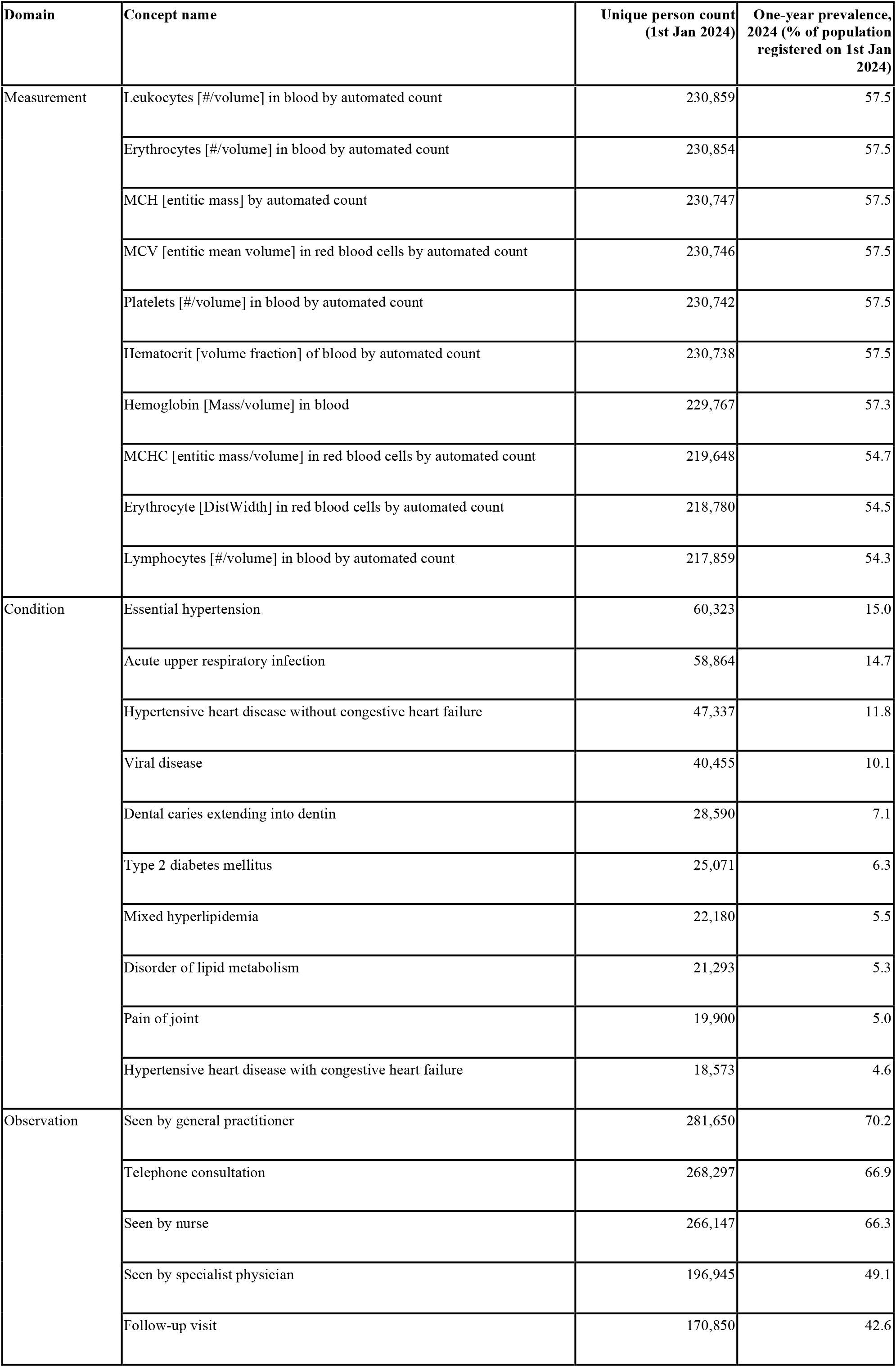
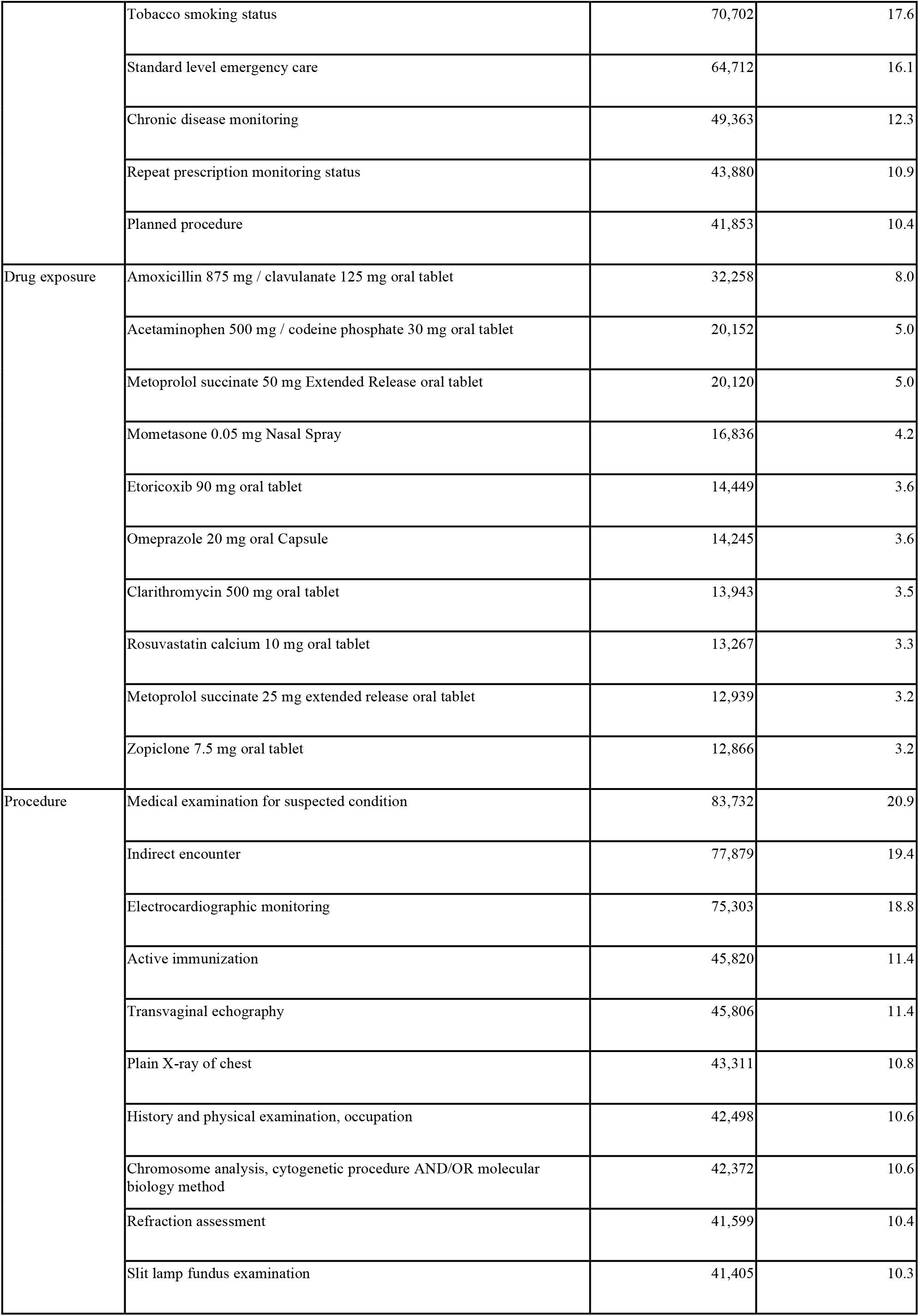
Ten most prevalent concepts per domain in EST-Health-30 (OMOP CDM)

| Domain | Concept name | Unique person count<br>(1st Jan 2024) | One-year prevalence,<br>2024 (% of population<br>registered on 1st Jan<br>2024) |
| --- | --- | --- | --- |
| Measurement | Leukocytes [# /volume] in blood by automated count | 230,859 | 57.5 |
|  | Erythrocytes [# /volume] in blood by automated count | 230,854 | 57.5 |
|  | MCH [entitic mass] by automated count | 230,747 | 57.5 |
|  | MCV [entitic mean volume] in red blood cells by automated count | 230,746 | 57.5 |
|  | Platelets [# /volume] in blood by automated count | 230,742 | 57.5 |
|  | Hematocrit [volume fraction] of blood by automated count | 230,738 | 57.5 |
|  | Hemoglobin [Mass/volume] in blood | 229,767 | 57.3 |
|  | MCHC [entitic mass/volume] in red blood cells by automated count | 219,648 | 54.7 |
|  | Erythrocyte [DistWidth] in red blood cells by automated count | 218,780 | 54.5 |
|  | Lymphocytes [# /volume] in blood by automated count | 217,859 | 54.3 |
| Condition | Essential hypertension | 60,323 | 15.0 |
|  | Acute upper respiratory infection | 58,864 | 14.7 |
|  | Hypertensive heart disease without congestive heart failure | 47,337 | 11.8 |
|  | Viral disease | 40,455 | 10.1 |
|  | Dental caries extending into dentin | 28,590 | 7.1 |
|  | Type 2 diabetes mellitus | 25,071 | 6.3 |
|  | Mixed hyperlipidemia | 22,180 | 5.5 |
|  | Disorder of lipid metabolism | 21,293 | 5.3 |
|  | Pain of joint | 19,900 | 5.0 |
|  | Hypertensive heart disease with congestive heart failure | 18,573 | 4.6 |
| Observation | Seen by general practitioner | 281,650 | 70.2 |
|  | Telephone consultation | 268,297 | 66.9 |
|  | Seen by nurse | 266,147 | 66.3 |
|  | Seen by specialist physician | 196,945 | 49.1 |
|  | Follow-up visit | 170,850 | 42.6 |
|  | Tobacco smoking status | 70,702 | 17.6 |
|  | Standard level emergency care | 64,712 | 16.1 |
|  | Chronic disease monitoring | 49,363 | 12.3 |
|  | Repeat prescription monitoring status | 43,880 | 10.9 |
|  | Planned procedure | 41,853 | 10.4 |
| Drug exposure | Amoxicillin 875 mg / clavulanate 125 mg oral tablet | 32,258 | 8.0 |
|  | Acetaminophen 500 mg / codeine phosphate 30 mg oral tablet | 20,152 | 5.0 |
|  | Metoprolol succinate 50 mg Extended Release oral tablet | 20,120 | 5.0 |
|  | Mometasone 0.05 mg Nasal Spray | 16,836 | 4.2 |
|  | Etoricoxib 90 mg oral tablet | 14,449 | 3.6 |
|  | Omeprazole 20 mg oral Capsule | 14,245 | 3.6 |
|  | Clarithromycin 500 mg oral tablet | 13,943 | 3.5 |
|  | Rosuvastatin calcium 10 mg oral tablet | 13,267 | 3.3 |
|  | Metoprolol succinate 25 mg extended release oral tablet | 12,939 | 3.2 |
|  | Zopiclone 7.5 mg oral tablet | 12,866 | 3.2 |
| Procedure | Medical examination for suspected condition | 83,732 | 20.9 |
|  | Indirect encounter | 77,879 | 19.4 |
|  | Electrocardiographic monitoring | 75,303 | 18.8 |
|  | Active immunization | 45,820 | 11.4 |
|  | Transvaginal echography | 45,806 | 11.4 |
|  | Plain X-ray of chest | 43,311 | 10.8 |
|  | History and physical examination, occupation | 42,498 | 10.6 |
|  | Chromosome analysis, cytogenetic procedure AND/OR molecular biology method | 42,372 | 10.6 |
|  | Refraction assessment | 41,599 | 10.4 |
|  | Slit lamp fundus examination | 41,405 | 10.3 |

## Data quality

Data cleaning, standardisation, and quality assessment were performed as part of the OMOP transformation pipeline, including validation, deduplication, and consistency checks, as described in Oja et al.^6^ Standardised data quality metrics were evaluated across all source datasets and OMOP domains to ensure completeness, consistency, and plausibility prior to release. This approach is aligned with the principles of the European Medicines Agency (EMA) Data Quality Framework^8^ for real-world data, which emphasises structured metrics, transparency of data provenance, and assessment of fitness for use in relation to a research question.

We additionally used the CdmOnboarding package ^9^ to generate an onboarding report of the type used by the DARWIN EU Coordination Centre and EMA to assess CDM readiness for regulatory studies. The report provides a comprehensive overview of all domains, including fill rates and mapping completeness. Embedded within the package, DataQualityDashboard ^10^ executed 2,374 checks covering plausibility, conformance, and completeness. Of 1,625 applicable checks, 1,612 passed (99.2%). The 13 failed checks were reviewed manually and found to reflect expected data characteristics that do not compromise data quality.

To assess population representativeness, we compared the age–sex distribution of individuals alive on 1 January 2024 with official Statistics Estonia population figures for the same date (**Figure 1**). The population size in EST-Health-30 is slightly below what would be expected from official Estonian population statistics (29.2% rather than 30.0%), particularly in the 20–34 age group. We attribute this primarily to recent immigrants who have no health insurance and have had no contact with the Estonian healthcare system.

This study follows the REporting of studies Conducted using Observational Routinely-collected Data (RECORD) statement ^11^. A completed RECORD checklist is provided as **Supplementary Table S1**.

## Secure processing environment

All data processing is carried out in the Secure Analysis and Processing Unit (SAPU), a secure processing environment hosted at the High Performance Computing Centre of the University of Tartu ^12^, which is fully compliant with the General Data Protection Regulation (GDPR) and European Health Data Space (EHDS) cybersecurity standards. These environments enable authorized researchers to process pseudonymised or anonymised health data without it leaving the controlled infrastructure. SAPU operates in a firewalled, internet-isolated environment, ensuring that data never leaves the secure perimeter; only aggregated results may be exported through a controlled disclosure procedure. All user activity is logged and recorded via continuous screen capture. In addition, SAPU is equipped with graphics processing unit (GPU) infrastructure, enabling the use of transformer-based models, including large language models such as Google MedGemma ^13^, within the SAPU environment.

## Data resource use

EST-Health-30 supports a wide range of real-world evidence (RWE) studies through its population coverage, longitudinal depth, and standardized OMOP structure. A key capability is the construction of detailed cohorts based on multimodal patient data.

For example, a heart failure cohort can be defined by combining diagnostic information, prescriptions, and laboratory measurements. Patients without prior heart failure diagnoses or related medication use can be identified at first presentation using indicators such as elevated NT-proBNP (≥125 ng/L) together with reduced ejection fraction (<40%). This illustrates the ability of the dataset to define clinically meaningful cohorts using structured variables across multiple data domains.

At the population level, the dataset enables characterisation of disease burden across the life course. **Figure 2** presents age-specific counts of distinct diagnoses per person stratified by ICD-10 chapters (used here for descriptive aggregation), demonstrating the shift from infectious and respiratory conditions in early life to chronic, cardiovascular, and metabolic diseases at older ages. A noticeable decline in diagnoses from the digestive system category in early adulthood reflects the transition at age 19 when dental care is no longer routinely covered by the health insurance system, leading to fewer recorded diagnoses in this domain. These patterns illustrate the ability of the dataset to capture population-level morbidity and its evolution over time, while also highlighting how healthcare system characteristics influence observed data.

**Figure 2.**
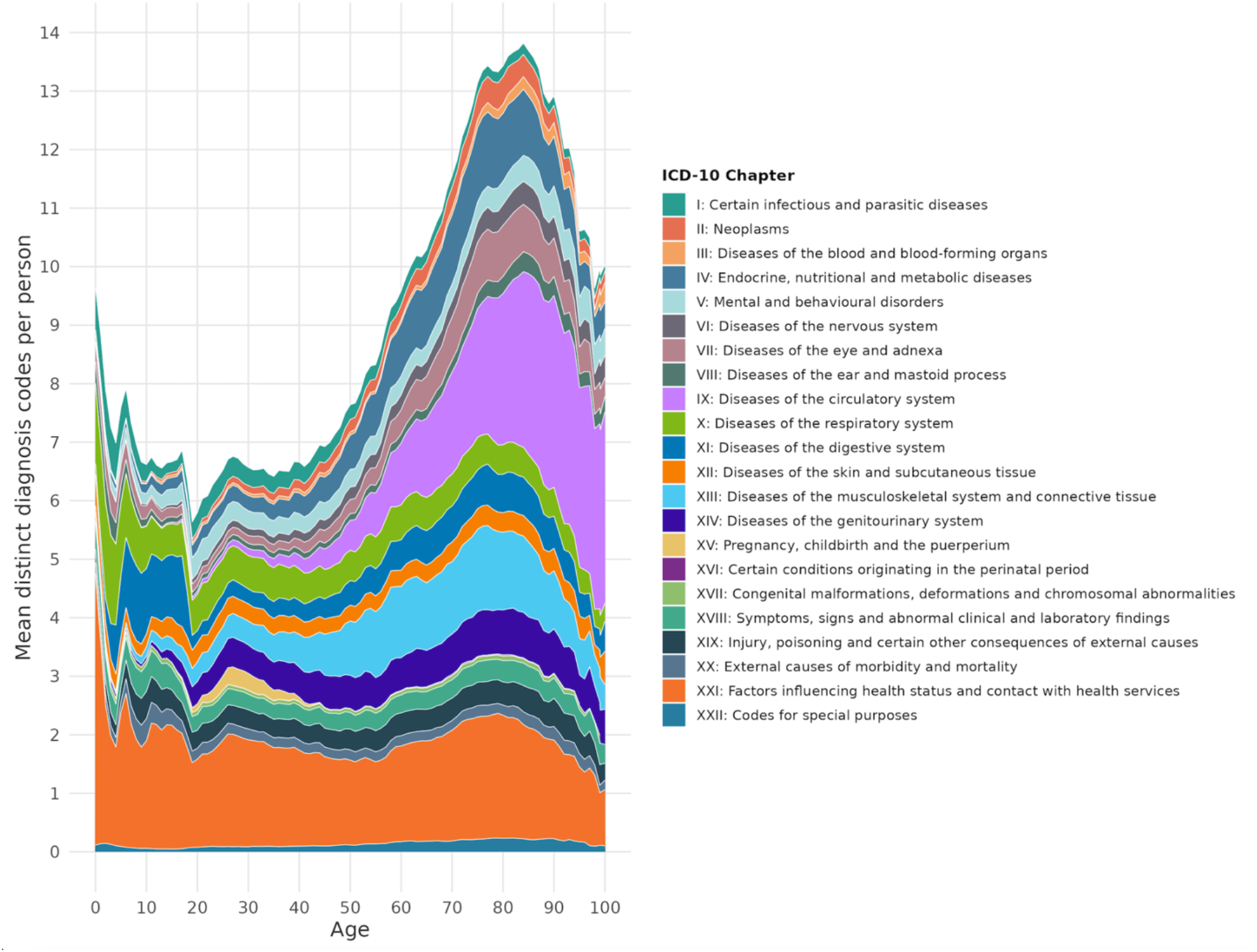
Age-specific distribution of diagnoses in 2024 by ICD-10 chapter. Values represent the mean number of distinct diagnoses per person at each age within each chapter. Estimates are smoothed using a 2-year rolling window.

The longitudinal structure further supports studies of disease trajectories, healthcare utilisation, healthcare costs, medication adherence, and adherence to clinical guidelines. Comparative effectiveness and safety analyses are possible using established causal inference methods, and detailed patient histories enable development and validation of predictive models for outcomes such as disease onset, hospitalisation, or mortality.

In addition to traditional epidemiological analyses, the dataset supports methodological research on data standardisation, phenotyping, and reproducible analytics. Its transformation to the OMOP CDM and integration of multiple healthcare data sources provide a practical setting for developing and evaluating cohort definitions, feature engineering pipelines, and reusable analytical workflows.

Finally, mapping to the OMOP CDM enables participation in international federated studies, where analyses are executed locally using shared analytical protocols and only aggregated results are exchanged. This allows EST-Health-30 to contribute to large-scale, multi-country studies while maintaining data privacy and supporting reproducible research.

## Strengths and weaknesses

EST-Health-30 covers a random 30% of the Estonian population with demographic distributions closely aligned with national statistics, supporting generalisability and reducing selection bias. Deterministic linkage across five nationwide databases enables comprehensive longitudinal capture of healthcare trajectories across care settings over more than a decade. The dataset provides rich multimodal clinical information — diagnoses, laboratory measurements, procedures, medications, and healthcare utilisation — standardised to the OMOP CDM using international vocabularies, enhancing interoperability and supporting federated research.

While EST-Health-30 provides rich longitudinal data, several limitations should be considered. As with all routinely collected health data, the source data were not originally created for research purposes, and may therefore be subject to misclassification, incomplete recording, and unmeasured confounding. The dataset does not capture periods during which individuals may have resided outside Estonia, which may lead to incomplete follow-up for some individuals. Although EHRs are included, claims and costs for services fully paid out of pocket or provided in private settings are not recorded. Dentistry (except for claims data for children), medical imaging data, and in-hospital medication use are also not captured.

Some important clinical information is recorded only in free-text fields and may not yet be fully available for analysis, as extraction of these data is ongoing. De-pseudonymisation of individuals is not possible, and caution is required when combining EST-Health-30 with other Estonian datasets due to potential patient overlap. In addition, relationships between individuals are not recorded, limiting family-level analyses.

The dataset currently contains limited direct information on socioeconomic factors such as education, income, or occupation. Although proxy indicators, such as insurance status and healthcare utilisation patterns, may be used, the lack of comprehensive socioeconomic data should be considered when addressing confounding in epidemiological studies.

Finally, despite including nearly 510,000 individuals, the dataset may still have limited statistical power for analyses of rare outcomes.

## Data resource access

EST-Health-30 was developed in accordance with the FAIR data principles. The dataset will be made findable upon publication of this manuscript. Access is governed through a multi-step process requiring a cooperation agreement with the Health Informatics Research Group at the University of Tartu, ethics approval from the national scientific ethics committee coordinated by the Estonian Research Council, and subsequent authorization from the Ministry of Social Affairs. Following approval, a minimal dataset is provided via SAPU, from which only aggregated results may be exported. The process from study protocol submission to data access typically takes approximately four to five months. Interoperability is achieved through standardization to the OMOP Common Data Model (CDM), enabling reuse of analytical tools, phenotype libraries, and federated network study protocols developed within the OHDSI community.Reusability is supported by comprehensive metadata, a detailed data dictionary, and harmonisation documentation as described in Oja et al^6^.

## Ethics approval

The compilation and use of the EST-Health-30 dataset was approved by the Estonian Bioethics and Human Research Council (no. 1.1-12/817) on 3 March 2025.

## Author contributions

Conceptualisation: SR, JV, RK, MO, SL;

Data curation: MO, ST, AS, HAT;

Formal analysis: SR;

Funding acquisition: JV, RK;

Investigation: SR;

Methodology: SR;

Resources: JV;

Validation: MO, AS;

Visualisation: SR;

Writing – original draft: SR;

Writing – review & editing: all authors.

All authors contributed to the interpretation of the data, critically revised the manuscript and approved the final version.

## Supplementary data

Supplementary data are available at IJE online.

## Use of artificial intelligence (AI) tools

During the preparation of this work, the authors used ChatGPT and Claude to improve the language, readability, and wording of the manuscript. After using this tool, the authors reviewed and edited the content as needed and take full responsibility for the accuracy, integrity, and final version of the publication.

## Data Availability

Access is governed through a multi-step process requiring a cooperation agreement with the Health Informatics Research Group at the University of Tartu, ethics approval from the national scientific ethics committee coordinated by the Estonian Research Council, and subsequent authorization from the Ministry of Social Affairs.

